# Development and Feasibility Evaluation of a Telehealth System (Impak Sihat) to Empower Rural Population in Malaysia on the Quality Use of Medicines

**DOI:** 10.1101/2024.12.22.24319506

**Authors:** Nor Ilyani Mohamed Nazar, Norny Syafinaz Ab Rahman, Nor Elina Alias, Syahrir Zaini, Tg Karmila Tg Mohd Kamil, Nurjasmine Aida Jamani, Mohamed Hassan Elnaem

**Affiliations:** Kulliyyah of Pharmacy, International Islamic University Malaysia (IIUM); Kulliyyah of Medicine, International Islamic University Malaysia (IIUM); School of Pharmacy and Pharmaceutical Sciences, Ulster University, Coleraine, United Kingdom

**Keywords:** Telehealth system, rural population, Malaysia, Quality Use of Medicines

## Abstract

The global rise of chronic diseases necessitates quality use of medicines (QUM) and adherence for effective management. Insufficient QUM understanding among patients adversely affects healthcare outcomes in Malaysia. High rates of irrational medication use, particularly in rural areas, are influenced by various factors. Continuous engagement from healthcare providers is essential, potentially facilitated by a tailored telehealth system. This study aimed to design a customized telehealth system for Malaysia’s rural population and assess patient experiences and acceptance. The study comprised three phases: i) a qualitative pre-development phase exploring internet access and health information-seeking behaviours; ii)development of the telehealth system (Impak Sihat-healthy impact) based on identified needs; and iii) a post-development feasibility evaluation of the Impak Sihat system through community demonstration and validated questionnaires. Fifteen respondents were interviewed during the pre-development phase, achieving saturation and revealing insights on health issues, internet usage, social media preferences, and technology literacy. A customized telehealth system was developed, addressing the identified needs with relevant content and resources. Following system development, a feasibility study with 77 participants indicated acceptable scores of 73 to 87% across six domains. However, older age was significantly linked to lower scores in the ‘User-friendly features and ease of learning and understanding’ domain (p<0.01). The telehealth system effectively meets Malaysia’s rural population’s needs and has been successfully developed. To optimize this system’s effectiveness, considerations regarding older age, socioeconomic status, internet reliability, and information confidentiality must be addressed.

## 1. Introduction

Quality Use of Medicines (QUM) encompasses the effective use and management of medications in treating chronic illnesses and maintaining overall health, considering the perspectives of healthcare practitioners, systems, communities, and patients. QUM emphasizes the careful selection, usage, management, and monitoring of medications to encourage rational drug use, thereby minimizing misuse, overuse, and underuse. Additionally, it seeks to empower individuals to address issues related to medications, including side effects and the complexities of managing multiple prescriptions [1]. Effective QUM management by pharmacists and other healthcare practitioners could positively affect patients’ clinical outcomes and improve the healthcare system’s organization [2]. In Asian countries such as Malaysia, the rate of medication nonadherence was reported to be around 50%–80% among patients with chronic illnesses [3]. Several studies suggest that the rural population has contributed to the high rate of nonadherence compared to the urban population [4].

Digital or telehealth refers to using information and communications technologies in medicine and other health professions to manage illnesses and health risks and promote wellness in the community [5]. Various innovations and applications of these technologies have been established in many developed countries. However, its application to improve populations’ health and well-being still has a large gap, especially in developing countries [6]. The recent COVID-19 pandemic has no doubt re-emerged the utilization of telehealth as part of an integral platform to reach out to the community in need during the global lockdown [7]. In Malaysia, The telemedicine blueprint was evidence of the government’s commitment to harnessing the potential of information and communication technology (ICT) to promote more equitable access to healthcare services for rural and remote areas [8]. Digital Health Malaysia (DHM), previously known as the Telemedicine Development Group (TDG), was formed in 2015 following the first telemedicine conference [9]. It is a platform where healthcare professionals, researchers, and the industry collaborate to advance the digital health agenda. However, despite its perceived advantages, digital health initiatives and services have not successfully reached and benefited the rural population of this country [8,10]. A study on healthcare team readiness for telehealth services in Malaysia revealed suboptimal results where two-thirds believed new technology could enhance current practices, while approximately half of the participants were willing to use telemedicine [11].

Telehealth provides key benefits for remote and rural areas with limited healthcare access. Utilizing health information technology improves pharmacist accessibility, enhances patient satisfaction and quality of life, and achieves better clinical outcomes while reducing resource use [12]. Telehealth modalities include telephonic outreach and specialized tools designed to increase health literacy. eHealth and telehealth medication adherence interventions were associated with improved medication possession ratio (MPR) and/or proportion of days covered (PDC) rates [13]. Telehealth models were associated with positive outcomes for patients and healthcare professionals, suggesting these models are feasible and can be effective. Future telehealth interventions and studies examining these programs are warranted, especially in rural communities, and future research in this population is highly recommended [14]. Engaging and empowering rural communities through health education and telehealth services is essential. These services can significantly reduce medication errors and adverse drug events while fostering long-term sustainability. However, current telehealth systems often fail to address the specific needs of rural populations. This study aims to develop a digital health system tailored to the understanding levels of rural residents and to evaluate their experiences with the new system, IMPAK SIHAT, which translates to “healthy impact” in the local language.

## 2. Methodology

The study has been registered and approved by the IIUM Research Ethics Committee (IREC 2024-194). Participation in all phases was entirely voluntary, and informed consent was sought from all willing to participate. The study was divided into three (3) main phases, and different study designs were applied. The study phases are detailed below: -

### 2.1. Qualitative pre-development phase exploring internet access and health information-seeking behaviours

Fifteen participants from the local community were recruited via a convenient sampling method and interviewed using an open-ended survey to explore the community’s internet accessibility or issues and the use of the internet or social media for searching for health-related information. The findings have reached saturation with 15 participants. The responses were coded and analyzed via thematic analysis. Significant themes were considered and integrated into the development of the telehealth system.

### 2.2. Development of the telehealth system (Impak Sihat) based on identified needs

The telehealth system was developed by a local system development vendor with a healthcare background, vast experience, and familiarity with the healthcare system data. A research assistant was appointed to create educational materials (videos/posters/infographics) to be incorporated into the system. The team members/experts carefully reviewed the materials to ensure their usability among rural communities. The system was tailor-made to suit the needs of the rural population in Malaysia with simplified, lay language and user-friendly features. A series of meetings and a system demonstration session by the vendor to the research team were regularly conducted to ensure that the system was congruent with the expected needs of the teamr members and the rural population. The system incorporated special features on patients’ health education and appointment booking to ease the communication between patients and healthcare practitioners.

### 2.3. Post-development feasibility evaluation of the Impak Sihat system through community demonstration and validated questionnaires

Once the system was about 80% complete, a feasibility study was conducted via telehealth system demonstration directly to the potential users among the rural population. A questionnaire was adapted and translated from the Telehealth Usability Questionnaire (TUQ) [15]. Five experts among the healthcare practitioners and scholars in the field were invited to review the content validity of the translated version. The content validity index (CVI) rated by the five experts was 0.919, and the Cronbach alpha value was 0.917, which shows good validity and internal consistency of the questionnaire. The validated questionnaire was distributed to the participants for feedback on usefulness, reliability, learnability, user experience, and future use recommendations. The inclusion criteria for the participants are being more than 18 years old, signing the informed consent form, being able to read and write, and regularly using a smartphone. The participants were introduced to the system’s essential features, mainly health education, appointment booking, and live video calls with healthcare practitioners. They were taught how to search for health information using the keywords on their mobile devices, set their appointments, and experience actual video calls with healthcare practitioners.

## 3. Results

### 3.1. Key themes identified from the pre-development phase activity

#### 3.1.1. Availability of smartphone devices with inconsistent internet coverage

Most respondents owned a smartphone (Android) and used their data (4G) for internet browsing, which cost them between RM30 and 45 per month. However, the internet speed test and coverage could have been more consistent in different areas of the villages and with varying weather conditions. This finding provides insight into the later implementation of telehealth services in this rural area, whereby these high-speed internet areas should be further identified and gazetted as telehealth stations for the local community, especially for live streaming consultation.

#### 3.1.2. Active use of social media

Most respondents were active on social media, and many had more than one (1) account, mainly WhatsApp, Facebook, Instagram and TikTok apps. They spend 1 to 10 hours browsing social media daily, primarily for leisure purposes - music, recipes, news, politics, socializing, and online shopping. Only a few local communities use the internet for work-related matters since the community’s economic activity is related to small-scale agricultural activities. Those who spend over 5 hours daily admit they love watching short videos, especially from TikTok. This information provides the team with meaningful insight into further engaging with the local community via social media platforms that must be linked with our telehealth system.

#### 3.1.3. Health literacy and internet usage for health-related information

Health Literacy is the ability to access, understand, appraise, and use information to make healthier choices [16]. Six respondents from the local community admitted using the internet to find health information, such as symptoms of diseases, traditional medicine, massages and health tips. They needed clarification when further asked about the validity and reliability of the gained information. Still, most of the time, they tend to buy or try out any products/ recommendations/ advice/ tips they encounter on the internet. This is an important finding on the health literacy of this rural population and a vast opportunity for community education and empowerment from our side as the provider.

### 3.2. Telehealth system development

The developed telehealth system has two (2) important components: i) for the public and ii) for healthcare practitioners with login credentials. Public users do not need to register login credentials and can use the system immediately. Public users can freely browse and go through the educational materials on various health-related topics, including medication usage, disease prevention, and lifestyle management, which have been developed and integrated into the system as an educational portal. The developed materials were simplified, mainly using laymen’s terms and with more infographic designs. Another vital feature for public users is the ‘appointment-booking feature’. Through this feature, public users can make appointments and communicate with healthcare practitioners to ask about any health-related information to the healthcare practitioners in charge without the need to be physically present at the healthcare facilities.

The second component for healthcare practitioners is registering and uniquely creating their login credentials; they can only access patients’ data on disease progression, vital signs and laboratory investigations, medication prescriptions, dosage, and adherence. Security System Development for Patient Data Management is crucial to telehealth system development. The vendor has employed industry-standard security measures to safeguard patient data effectively. This includes implementing encryption techniques, access controls, and regular security audits to mitigate potential risks and vulnerabilities. Encryption techniques such as Transport Layer Security (TLS) have also ensured secure communication [17]. The content performance analysis feature has also been incorporated into the system. This feature can provide information to the administrators on the:-

i Most Accessed Materials: Showcase a list or heatmap of the most accessed educational materials and resources.
ii Popular Topics: Display a tag cloud or bar chart highlighting popular topics and categories among users.
iii Feedback and Ratings: Present a radar chart or stacked bar chart summarising feedback and ratings for content quality and relevance.
vi Content Consumption Patterns: Visualize a heatmap or line graph indicating content consumption patterns, including peak usage times and average session durations.

### 3.3. Feasibility study on user’s acceptance and experience

In this feasibility study, the local community participated in a demonstration session of the telehealth system, and their feedback was collected cross-sectionally via a validated questionnaire.

#### 3.3.1. System demonstration implementation

A telehealth system demonstration and feasibility evaluation were conducted at two rural villages of Ulu Tembeling, Kg. Kuala Sat and Kg. Bantal in the District of Jerantut Pahang. Seventy-seven patients who met the inclusion criteria consented to the telehealth demonstration session. Fifty-one (66.2%) respondents were female, and another 33.8% were male. The mean age was 53.4 (±11.8) years old. Most respondents were housewives (48.1%) for females and self-employed or working in agricultural sectors for males (39.0%). Table 1 lists the summary of demographic data collected.

**Table 1:**
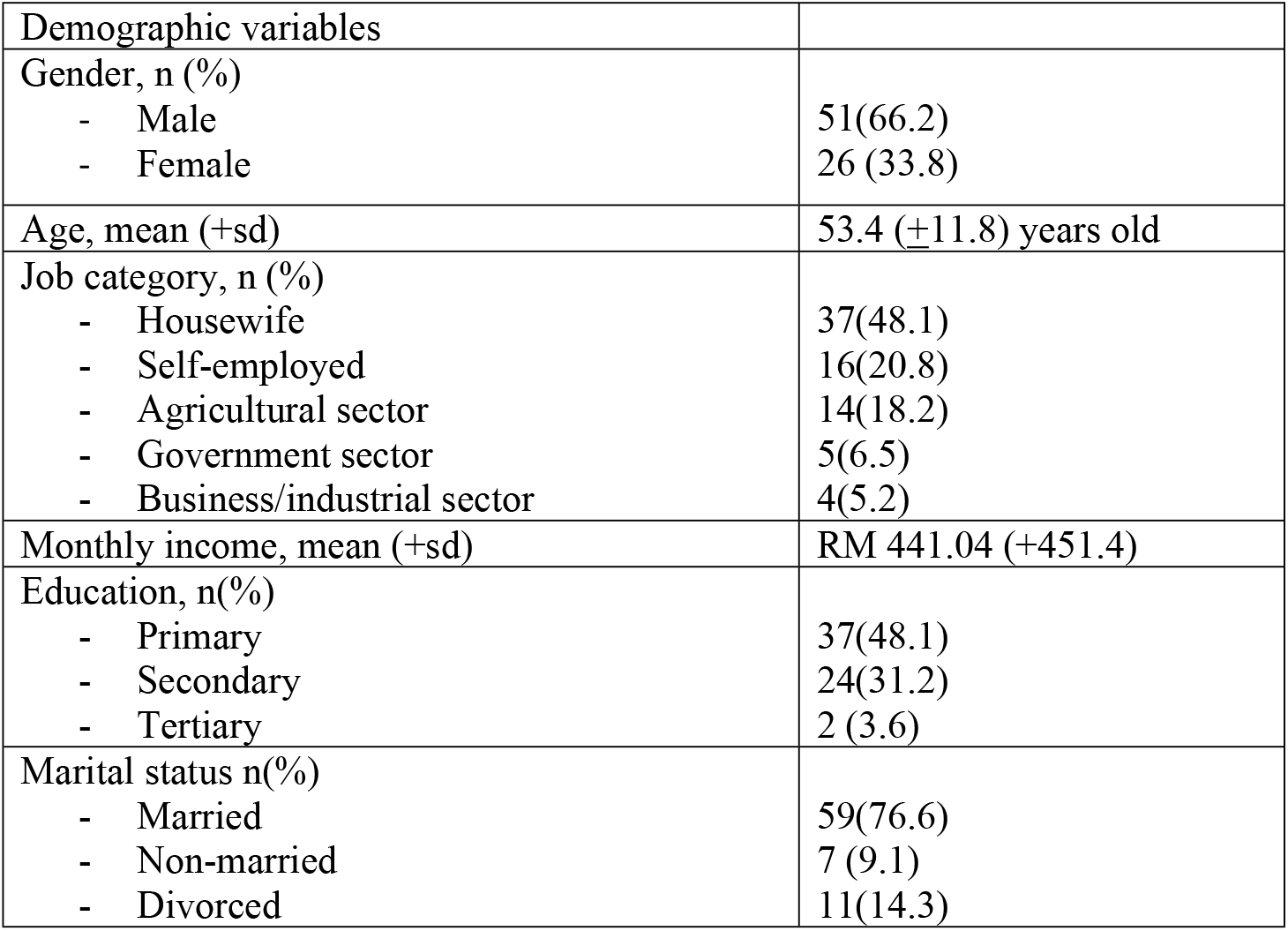
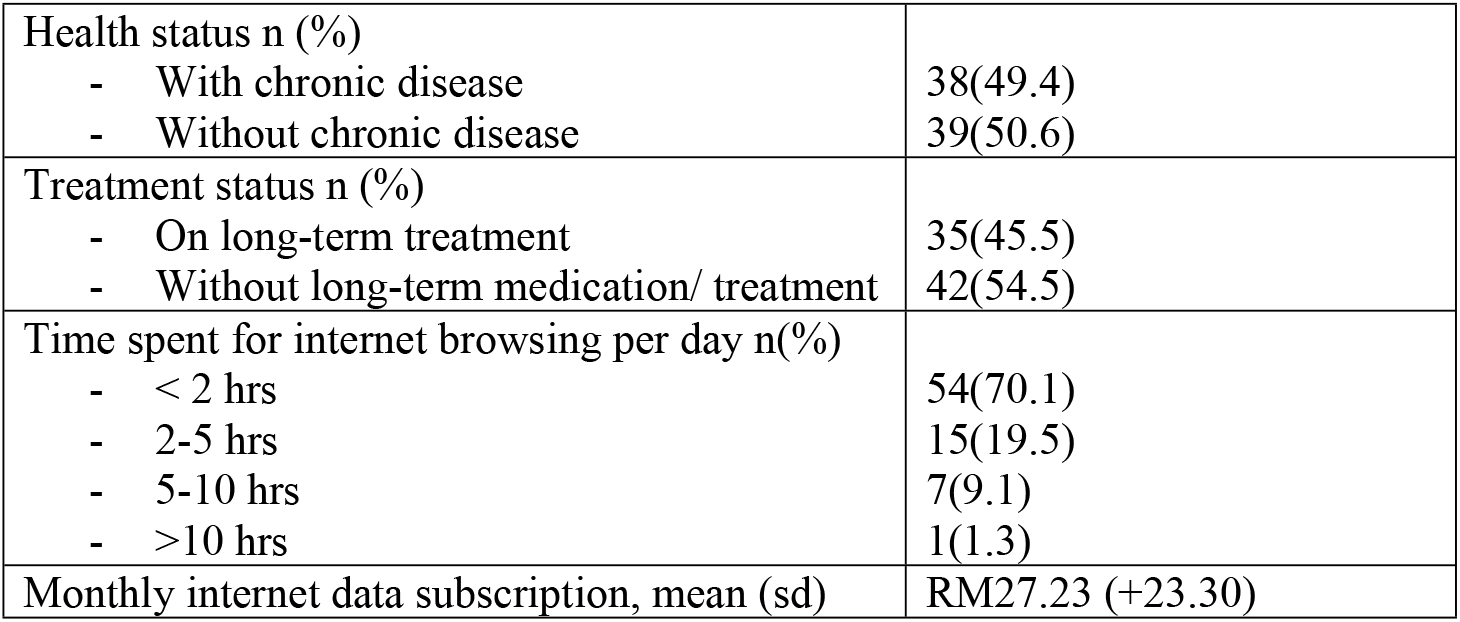
Demographic data of the local respondents who participated in the post-development study.

#### 3.3.2. Feasibility Evaluation

The feasibility evaluation of the telehealth system was translated into the index scores for all six domains measured as displayed in Table 2.

**Table 2:** Index scores for all six domains of the feasibility evaluation of the developed telehealth system.

| Domain | Description | Index score (%) |
| --- | --- | --- |
| 1 | The usefulness of the Telehealth System | 87.0 |
| 2 | User-friendly features and ease of learning and understanding. | 74.8 |
| 3 | Audiovisual quality of the system. | 75.3 |
| 4 | Interaction quality with healthcare practitioners. | 80.7 |
| 5 | Reliability of information retrieved from the system and data confidentiality. | 73.1 |
| 6 | Users'overall satisfaction. | 80.3 |

A significant negative correlation was found between the age and the scores for domain 2; the higher the age, the lower the scores for this domain (p<0.01). A poor to moderate positive correlation existed between the index scores for each domain (0.275-0.623, p<0.01). There was no significant correlation between the index scores and gender, monthly income, education level or job categories of the respondents. Even though there was a significant negative correlation (p<0.01) between patients’ health status with domains 2,5 and 6, the correlation was not strong (−0.227, -0.287 and -0.263, respectively).

## 4. Discussion

### 4.1. Community acceptance for rural telehealth initiatives

Health technology interventions are essential to rural healthcare services as they enhance residents’ access to care (including speciality care) that they would not otherwise be able to get in remote areas and reduce staffing costs, travel expenses, and travel time [14]. Rural residents either travel great distances to receive care, have limited access to healthcare, or are delayed in seeking care until after an emergency. Poor health outcomes and a financial and social burden on the patient and the healthcare system can arise from limited access to care. An additional strain is placed on the patient by the expense of travelling for medical care, including lost work hours, decreased productivity, and higher childcare or caregiver support [18]. Telehealth extends the reach of health services and provides the opportunity to reduce barriers to care in rural communities.

This study in a rural community in Malaysia revealed an overall positive acceptance and experience among the local community towards the newly developed telehealth system – Impak Sihat, which aims to provide health education and address the issues of chronic diseases and quality use of medicine among disease progression in the rural population. This is consistent with a similar study in a rural community, which found that 88% of respondents were either considering or favouring telehealth, and only 12% of respondents reported that they would not be open to telehealth [19]. Telehealth perceptions were generally favourable among rural patients and providers, although satisfaction was lower among older patients and providers. Another finding has strongly suggested that telehealth approaches may add value and efficiency to rural clinical practice [20].

### 4.2. Issues to consider before widespread adoption

Despite the positive and favourable acceptance of the current telehealth initiatives, several issues need to be addressed in our local rural setting before this initiative can be fully implemented.

#### 4.2.1. Aging population of the rural community

The mean age of participants in this current study is 53 (±11.8) years old. The result also demonstrated a strong correlation between the age with user-friendly features and the ease of learning and understanding (p<0.05), where the lower the age, the higher the index score for this domain. This is expected as many of those shown interest in using telehealth were from the younger generation [19]. A study in a rural population with 206 respondents (mean age was 60) found that video conferencing and patient portal use pose barriers to older people with less education. However, these barriers disappear when telehealth is available through the telephone. Being younger, married/partnered, and having some college education were significantly associated with patient portal use [21].

#### 4.2.2. Low socioeconomic status (SES)

Low SES in rural Malaysia is a pronounced issue that presents significant challenges for individuals and communities. Despite Malaysia’s rapid economic growth and urbanization, rural areas struggle with poverty, limited access to education and healthcare, and inadequate infrastructure. This study shows that most of the rural population has low socioeconomic status, which might affect their ability to acquire additional cost of internet data if the telehealth service is provided to them. Hence, it should be designed as a web-based or with minimal data consumption if it is to be developed and implemented as a mobile application. Studies found that increasing age and being in the lowest median household income quartile were associated with lower odds of completing virtual visits overall [22,23].

#### 4.2.3. Unstable internet access in the rural area

Malaysia still needs help with unstable internet access, especially in rural areas. One of the central themes from the qualitative findings of the current study is unstable internet access in an area that heavily depends on local weather. This remains a significant challenge that might hamper telehealth development and exacerbate healthcare disparities between urban and rural communities.

While urban regions in Malaysia enjoy high-speed internet connectivity of more than 90% coverage and advanced digital infrastructure, rural areas often struggle with inadequate internet services (<70% coverage) and poor facilities [24]. Rural areas are often located in remote regions that are difficult to reach, making it costly and logistically challenging for service providers to deploy the necessary infrastructure [25]. Additionally, the lower population density in these areas does not present a financially viable market for telecommunications companies, leading to a lack of competition and innovation [25].

Therefore, cutting-edge options like satellite internet and neighbourhood Wi-Fi projects can be explored. In remote locations where traditional infrastructure is impracticable, satellite technology may be able to reach people [26]. Community-driven initiatives can empower rural populations and encourage local ownership of digital resources by bringing local communities together to create Internet services [25]. Furthermore, digital literacy programs should be implemented to equip rural residents with the skills needed to navigate the online world effectively. By fostering digital skills, individuals can better utilize available resources, engage in e-commerce, and access online education and healthcare services [27,28].

#### 4.2.4. Data confidentiality and trust issues

The index score for the system’s information reliability and data confidentiality domain shows the lowest score among the six domains, with a 73.1% score. Data confidentiality and reliability have become critical principles in the digital age since information is readily available, and the internet is a major source of knowledge. Data reliability refers to the accuracy, consistency, and trustworthiness of information available in the system. The fast growth of social media platforms has significantly impacted how individuals consume and interpret information. Hence, it is essential to ensure that the available content of the telehealth system is from valid and reliable resources.

While data reliability is a crucial component of the developed telehealth system, confidentiality is equally significant in shaping public perception. The internet has transformed the landscape of personal data collection, raising questions about how individuals’ information is stored, used, and protected. The public’s perception of internet data reliability and confidentiality has important societal ramifications [29]. A lack of trust in data confidentiality can lead to disengagement with the healthcare providers providing the telehealth service. Conversely, heightened awareness of data confidentiality issues can empower individuals to take control of their digital footprints. As users become more informed about their rights and the importance of data protection, they may demand greater accountability from corporations and advocate for stronger privacy regulations. This shift in perception can foster a culture of data responsibility, where both individuals and organizations prioritize ethical data practices.

## 5. Conclusion & recommendation

The advent of telehealth offers a promising solution to healthcare disparities and challenges in rural areas of Malaysia. The current study shows good acceptance of the developed telehealth system among the rural community participants. However, the system might need further modification and improvement before fully utilizing it in the rural community. Several issues also need to be considered and addressed prior to implementation, such as high rate of old age, low socioeconomic status, unstable internet access and information reliability/ confidentiality.

Unstable internet access in rural Malaysia is a pressing issue that requires urgent attention. The implications of this digital divide extend beyond mere connectivity; they affect education, healthcare, and economic opportunities for rural communities. By investing in infrastructure, exploring innovative technological solutions, and promoting digital literacy, Malaysia can work towards bridging the digital divide and ensuring that all its citizens, regardless of their geographical location, have equal access to the opportunities the internet offers. In doing so, Malaysia can foster inclusive growth and development, ultimately leading to a more equitable society.

## Data Availability

All relevant data are within the manuscript and its Supporting Information files.

N\A

## Acknowledgement

This work was supported by the ISPF Grant of the Networking Initiative for Digital Health System Development to Empower the Rural Population in Malaysia on the Quality Use of Medicines.

## Author Contributions

**Conceptualisation:** Nor ilyani Nazar, Mohamed Elnaem.

**Data curation:** Nor ilyani Nazar, Norny Syafinaz, Nor Elina Alias, Syahrir Zaini,Tg Karmila, and Nurjasmine

**Formal analysis:** Nor ilyani Nazar, Norny Syafinaz, Nor Elina Alias

**Funding acquisition:** Mohamed Elnaem

**Methodology:** Nor ilyani Nazar, Mohamed Elnaem.

**Writing – original draft:** Nor ilyani Nazar, Norny Syafinaz, Nor Elina Alias, Syahrir Zaini,Tg Karmila, and Nurjasmine

**Writing – review & editing:** Nor ilyani Nazar, Norny Syafinaz, Nor Elina Alias, Syahrir Zaini,Tg Karmila, Nurjasmine and Mohamed Elnaem

## References

1. National Medicines Policy. National Strategy for Quality Use of Medicines | Australian Government Department of Health and Aged Care. 2002. Available: https://www.health.gov.au/resources/publications/national-strategy-for-quality-use-of-medicines?language=en

2. Mohd-Tahir NA, Paraidathathu T, Li SC. Quality use of medicine in a developing economy: Measures to overcome challenges in the Malaysian healthcare system. SAGE Open Medicine. SAGE Publications Ltd; 2015. doi:10.1177/2050312115596864

3. Ganasegeran K, Rashid A. The prevalence of medication nonadherence in post-myocardial infarction survivors and its perceived barriers and psychological correlates: A cross-sectional study in a cardiac health facility in Malaysia. Patient Prefer Adherence. 2017;11: 1975–1985. doi:10.2147/PPA.S151053

4. Magnabosco P, Teraoka EC, De Oliveira EM, Felipe EA, Freitas D, Marchi-Alves LM. Comparative analysis of nonadherence to medication treatment for systemic arterial hypertension in urban and rural populations. Rev Lat Am Enfermagem. 2015;23: 20–27. doi:10.1590/0104-1169.0144.2520

5. WHO. Global Strategy on Digital Health 2020-2025. World Health Organization; 2021.

6. Spanakis EG, Santana S, Tsiknakis M, Marias K, Sakkalis V, Teixeira A, et al. Technology-based innovations to foster personalized healthy lifestyles and well-being:a targeted review. J Med Internet Res. 2016;18. doi:10.2196/jmir.4863

7. Elnaem MH, Nuffer W. Diabetes care and prevention services provided by pharmacists: Progress made during the COVID-19 pandemic and the need for additional efforts in the post-pandemic era. Exploratory Research in Clinical and Social Pharmacy. Elsevier Inc.; 2022. doi:10.1016/j.rcsop.2022.100137

8. Marzuki NM, Ismail S, Al-Sadat N, Mohsein A, Ehsan FZ. Evaluation of Telehealth implementation in government primary health clinics-A study protocol.

9. Homepage - Digital Health Malaysia. [cited 16 Dec 2024]. Available: https://www.digitalhealthmalaysia.org/

10. Speyer R, Denman D, Hons B, Wilkes-Gillan S, Chen YW, Bogaardt H, et al. Effects of telehealth by allied health professionals and nurses in rural and remote areas: A systematic review and meta-Analysis. Journal of Rehabilitation Medicine. Foundation for Rehabilitation Information; 2018. pp. 225–235. doi:10.2340/16501977-2297

11. Ang WC, Khalid K, Hat SZC; Anuar A. Evaluating Telemedicine Perception and Readiness among Healthcare Workers in Malaysia. Perspect Health Inf Manag. 2023;20.

12. Tegegne MD, Wubante SM, Melaku MS, Mengiste ND, Fentahun A, Zemene W, et al. Tele-pharmacy perception, knowledge and associated factors among pharmacy students in northwest Ethiopia: an input for implementers. BMC Med Educ. 2023;23. doi:10.1186/s12909-023-04111-9

13. Bingham JM, Black M, Anderson EJ, Li Y, Toselli N, Fox S, et al. Impact of Telehealth Interventions on Medication Adherence for Patients With Type 2 Diabetes, Hypertension, and/or Dyslipidemia: A Systematic Review. Ann Pharmacother. 2021;55: 637–649. doi:10.1177/1060028020950726

14. Butzner M, Cuffee Y. Telehealth interventions and outcomes across rural communities in the United States: Narrative review. J Med Internet Res. 2021;23: e29575. doi:10.2196/29575

15. Parmanto B, Lewis, Jr. AN, Graham KM, Bertolet MH. Development of the Telehealth Usability Questionnaire (TUQ). Int J Telerehabil. 2016;8: 3. doi:10.5195/IJT.2016.6196

16. Nutbeam D. The evolving concept of health literacy. Soc Sci Med. 2008;67: 2072– 2078. doi:10.1016/J.SOCSCIMED.2008.09.050

17. Hazra R, Chatterjee P, Singh Y, Podder G, Das T. Data Encryption and Secure Communication Protocols. https://services.igi-global.com/resolvedoi/resolve.aspx?doi=104018/979-8-3693-6557-1.ch022. 1AD; 546–570. doi:10.4018/979-8-3693-6557-1.CH022

18. Orlando JF, Beard M, Kumar S. Systematic review of patient and caregivers’ satisfaction with telehealth videoconferencing as a mode of service delivery in managing patients’ health. PLoS One. 2019;14. doi:10.1371/journal.pone.0221848

19. Kolluri S, Stead TS, Mangal RK, Lane Coffee R, Littell J, Ganti L. Telehealth in Response to the Rural Health Disparity. Health Psychol Res. 2022;10. doi:10.52965/001c.37445

20. Klee D, Pyne D, Kroll J, James W, Hirko KA. Rural patient and provider perceptions of telehealth implemented during the COVID-19 pandemic. BMC Health Serv Res. 2023;23. doi:10.1186/s12913-023-09994-4

21. Brunner W, Pullyblank K, Scribani M, Krupa N, Fink A, Kern M. Determinants of Telehealth Technologies in a Rural Population. Telemedicine and e-Health. 2023;29: 1530–1539. doi:10.1089/TMJ.2022.0490

22. Darrat I, Tam S, Boulis M, Williams AM. Socioeconomic Disparities in Patient Use of Telehealth during the Coronavirus Disease 2019 Surge. JAMA Otolaryngol Head Neck Surg. 2021;147: 287–295. doi:10.1001/jamaoto.2020.5161

23. Williams C, Shang D. Telehealth Usage Among Low-Income Racial and Ethnic Minority Populations During the COVID-19 Pandemic: Retrospective Observational Study. J Med Internet Res. 2023;25: e43604. doi:10.2196/43604

24. Tengku Mohamed FAZIHARUDEAN, Hitoshi MITOMO. Digital Divide among Public Servants in Malaysia: Urban-Rural Differences in Valuing the Use of the Internet. Studies in Regional Science. 2006;35: 837–849.

25. Alabdali SA, Pileggi SF, Cetindamar D. Influential Factors, Enablers, and Barriers to Adopting Smart Technology in Rural Regions: A Literature Review. Sustainability (Switzerland). MDPI; 2023. doi:10.3390/su15107908

26. Kamarudin S, Omar SZ, Bolong J, Osman MN, Mahamed M. ICT Development of Community in Rural Areas. International Journal of Academic Research in Business and Social Sciences. 2019;9. doi:10.6007/ijarbss/v9-i9/6273

27. Samsuddin SF, Mohamed NA, Bolong J. Understanding the Digital Capabilities of Rural Communities in Low Literacy Rate Areas in Malaysia towards Digital Society. International Journal of Academic Research in Business and Social Sciences. 2021;11. doi:10.6007/ijarbss/v11-i15/10644

28. Ji H, Dong J, Pan W, Yu Y. Associations between digital literacy, health literacy, and digital health behaviors among rural residents: evidence from Zhejiang, China. Int J Equity Health. 2024;23. doi:10.1186/s12939-024-02150-2

29. David A, Gomathi Sankar J, Azam S. INTERNET USERS TOP CONCERNS ENSURING DATA PRIVACY, SECURITY, AND PROTECTION. 10th International Conference on Multidisciplinary Research and Modern Education. 2023; 1–16. Available: https://doi.org/www.princetonpress.us

